# Separating representational and noise components of speech prosody perception after stroke

**DOI:** 10.1101/2023.10.17.23297140

**Authors:** Aynaz Adl Zarrabi, Mélissa Jeulin, Pauline Bardet, Pauline Commère, Lionel Naccache, JJ Aucouturier, Emmanuel Ponsot, Marie Villain

## Abstract

After a right hemisphere stroke, more than half of the patients are impaired in their capacity to produce or comprehend speech prosody. Yet, and despite its social-cognitive consequences for patients, aprosodia following stroke has received scant attention. In this report, we introduce a novel, simple psychophysical procedure which, by combining systematic digital manipulations of speech stimuli and reverse-correlation analysis, allows estimating the internal sensory representations that subtend how individual patients perceive speech prosody, and the level of internal noise that govern behavioral variability in how patients apply these representations. Tested on a sample of N=22 right-hemisphere stroke survivors and N=12 healthy controls, the representation+noise model provides a promising alternative to the clinical gold standard for evaluating aprosodia (MEC): both parameters strongly associate with receptive, and not expressive, aprosodia measured by MEC within the patient group; they have better sensitivity than MEC for separating high-functioning patients from controls; and have good specificity with respect to non-prosody-related impairments of auditory attention and processing. Taken together, individual differences in either internal representation, internal noise, or both, paint a potent portrait of the variety of sensory/cognitive mechanisms that can explain impairments of prosody processing after stroke.

## Introduction

After a right hemisphere stroke, more than half of the patients present a communication disorder such as aprosodia^1–3^, the impossibility to produce or comprehend speech prosody - or the “melody” of speech. Despite the social-cognitive implications for patients of not being able to process e.g. emotional or attitudinal expressions^4^, aprosodia following stroke has received scant attention: first, existing assessment tools based on simple perceptive tasks are insufficiently sensitive; second, we lack mechanistic understanding of why patients perform poorly on such tasks and therefore lack practical therapeutic targets for their subsequent rehabilitation^5^.

When studying the neural mechanisms that relate physical stimuli to perception, the modern field of psychophysics has largely moved from simply measuring sensory thresholds and psychometric functions, and now provides a toolbox of techniques to measure and fit multi-staged models able to simulate participant behaviour^6^. Notably for the example of speech prosody, the psychophysical technique of reverse-correlation (or “classification images”)^7^ allows estimating, at the individual level, not only what sensory representations subtend the normal or abnormal perception of e.g. interrogative prosody^8^, but also “internal noise” parameters that capture aspects of behavioral variability that are of potential neurological relevance^9,10^. While the representation + noise model has a rich history in healthy participants, with or without peripheral hearing impairment^11, 12^, its use in participants with neurological or developmental disorders has received relatively little attention^13,14,15^. Here we show on a sample of N=22 right-hemisphere brain stroke survivors that such simple procedures promise to enrich the current clinical toolbox with more sensitive and informative markers of receptive aprosodia.

## Materials and methods

### Participants

N=22 brain stroke survivors (male:17; M=57 yo, SD=13.0), and N=12 age-matched controls (male: 6;M=59 yo, SD=17.6) took part in the study. All participants were in- or out-patients of the Physical Medicine & Rehabilitation Department, APHP Pitié-Salpêtrière Hospital in Paris, France, undergoing speech therapy for different deficits post-stroke like swallowing difficulties, neurovisual impairments, attentional impairments, neglect, and dysphasia etc. Patients included in the study (Table 1) had a history of supratentorial right-hemisphere ischaemic stroke, corroborated by clinical assessments NIH stroke scale (NIHSS; M=10.8) and brain MRI, and dating less than 1y (M=4 months) at the time of inclusion; were first-language French speakers; and had no disorders of wakefulness/consciousness, dementia, severe dysarthria, psychiatric antecedents (>2 months in-patient) or major visual or auditory impairment (> 40dB HL). Patients with language comprehension deficits (score < 10/15 on the BDAE instruction-following task) were excluded from the study. In addition, a group of N=12 controls matched in age, sex and degree of hearing loss was recruited via the INSEAD-Sorbonne Université Center for Behavioural Science, also in Paris, France.

**Table 1.** Patients and control demographics and clinical characteristics. N=22 right-hemisphere stroke survivors and N=12 age-matched controls took part in the study. Abbreviations used: MEC: Montréal Evaluation de la Communication; BDAE: Boston Diagnostic Aphasia Examination

|  |  | controls | patients |
| --- | --- | --- | --- |
| **n** |  | 7 | 22 |
| **Sex, n (%)** | f | 3 (42.9%) | 5 (22.7%) |
|  | m | 4 (57.1%) | 17 (77.3%) |
| **Age, median [Q1,Q3]** |  | 59.0 yo [52.5,71.5] | 60.5 yo [52.2,63.0] |
| **Month after stroke, median [Q1,Q3]** |  |  | 4.0 mo [1.0,5.0] |
| **Stroke type, n (%)** | HEM |  | 3 (33.3%) |
|  | ISCH |  | 6 (66.7%) |
| **NIH stroke scale (NIHSS), median [Q1,Q3]** |  |  | 10.0 [5.5,16.0] |
| **MEC Prosody Comprehension item, median [Q1,Q3]** |  |  | 9.0 [8.0,11.0] |
| **MEC Prosody Repetition item, median [Q1,Q3]** |  |  | 11.0 [10.0,12.0] |
| **MEC Total, median [Q1,Q3]** |  |  | 21.0 [18.2,22.8] |
| **BDAE command execution item, median [Q1,Q3]** |  |  | 14.0 [14.0,15.0] |
| **Audiogram left-ear, median dBHL at 1000Hz [Q1,Q3]** |  | 0.0 dBHL [0.0,15.0] | 20.0 dBHL [15.0,30.0] |
| **Audiogram right-ear, median dBHL at 1000Hz [Q1,Q3]** |  | 5.0 dBHL [0.0,20.0] | 15.0 dBHL [7.5,37.5] |
| **Vocal audiogram, median% detection at 40dB [Q1,Q3]** |  |  | 99.0 % [94.0,100.0] |

### Clinical assessment

Two subtests of the French version of the “Montréal Evaluation de la Communication” (MEC) protocol^16^ were administered to the patients to assess their linguistic prosody capacities (comprehension and repetition). The linguistic prosody comprehension subtest evaluated the ability to identify linguistic intonation patterns. This subtest consists of four semantically neutral simple sentences and each one is presented to the patient with three different intonations, for a total of 12 items. After listening to a sentence the patient is asked to select the correct intonation among the three different written options (interrogative, imperative or affirmative). The linguistic prosody repetition subtest examines the ability to verbally reproduce linguistic intonations. It is formed of the same four sentences as the comprehension task. The previously recorded stimuli are presented in random order. The patient is asked to repeat each sentence with the same intonation. The maximum score is 12 for both subtests.

In order to exclude patients with a significant hearing impairment from the study, patients were assessed using Lafon’s cochlear lists of monosyllabic words (List 2 and List 3)^17^. These were calibrated at an intensity of 40 decibels (dB) and played through headphones. Only patients who scored 80% or more on both lists were included. In addition, the Boston Diagnostic Aphasia Examination (BDAE) command execution subtest^18^ was used to exclude patients with comprehension disorders. Only patients with a score of 12/15 or higher were included.

To assess possible mood disorders, the Hospital Anxiety and Depression Scale (HADS) self-questionnaire^19^ was administered to patients to assess their current level of anxiety and depression. It contains 7 questions for the anxiety part and 7 questions for the depression part, with a separate score for each. A score of 11 or more for each part indicates a possible anxiety and/or depression state.

To assess auditory attention, we used the sustained auditory attention subtest of the “Logiciel d’Attention en Modalité Auditive” (LAMA)^20^. The assessment and rehabilitation software “Aide Informatisée pour la Rééducation des Troubles Auditifs Centraux” (Airtac2)^21^ was used to assess central auditory processing. Intensity discrimination and duration discrimination of non-verbal sounds were proposed to compare central auditory processing abilities with the results of the Reverse Correlation task. Finally, the Montreal Battery of Evaluation of Amusia (MBEA)^22^ was selected to assess the music perception abilities of patients. Since the disorder of music perception (Amusia) is primarily a disorder of pitch perception^23^, the three tasks in the melodic organization part were selected.

### Procedure

We recorded a 426-ms utterance of the French word “vraiment” (“really”), and generated prosodic variations by dividing it into six segments of 71 ms and randomly manipulating the pitch of each breakpoint independently using a normal distribution (SD = 70 cents; clipped at ± 2.2 SD), hereafter referred to as “stimulus noise”. These values were linearly interpolated between time points and fed to an open-source pitch-shifting toolbox developed for this purpose^24^. We then presented patients with 150 successive pairs (controls: 600 pairs) of such manipulated utterances (*really/really?*) asking them to judge which, within each pair, sounded most interrogative. The sequence was divided into 3 blocks of 50 pairs for patients and 6 blocks of 100 pairs for controls. Without the participant’s knowing, the first and last block of each sequence contained identical pairs of sounds (a procedure called double-pass^9,25^, allowing us to examine response variability), but all other sounds in the sequence were otherwise distinct, N=9 patients were additionally tested 4 times, and these data points are not included in the statistics except for figure 3. Sounds were delivered using closed headphones (Beyerdynamics DT770) at a comfortable level (70 dB SPL). The inter-stimulus interval in each pair was 500 ms, and the interval between successive pairs was 1s. The procedure took about 15min to complete for patients and 1hr for controls.

**Figure 1.**
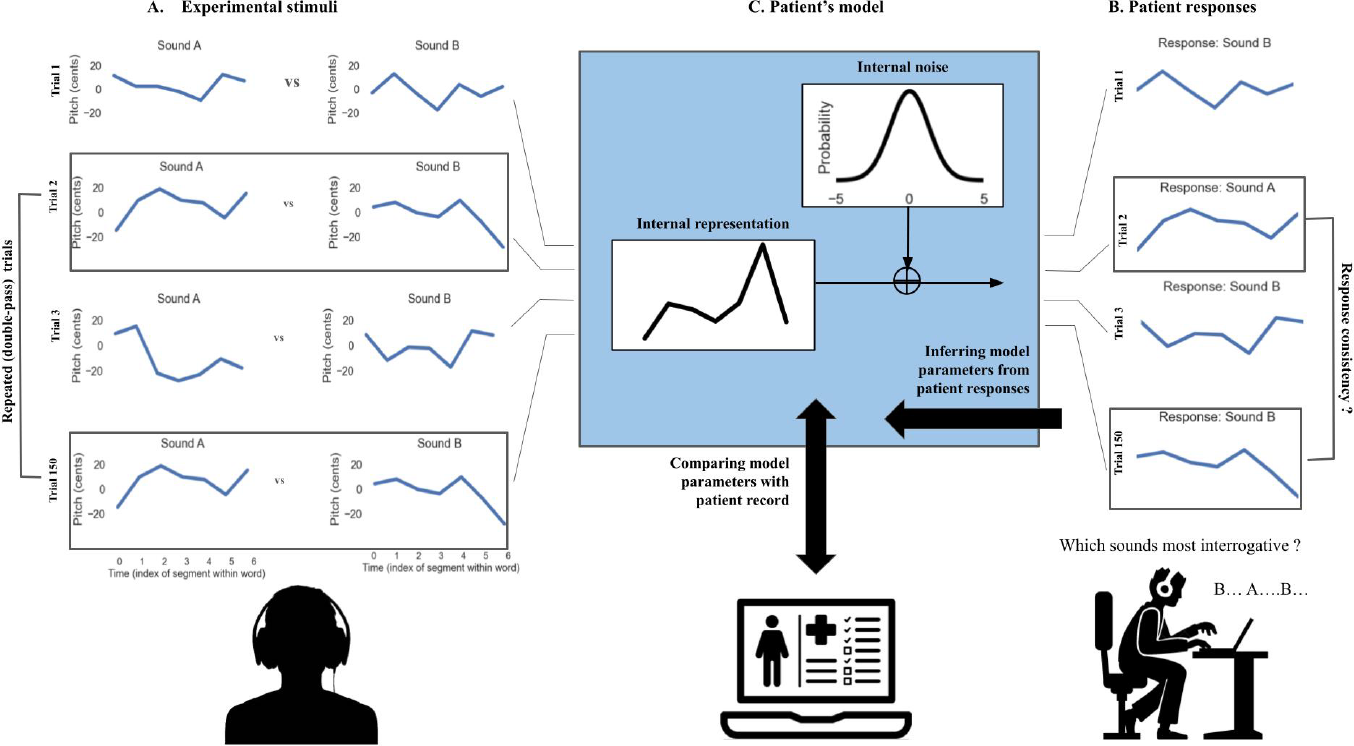
The representation + noise model. Patients were presented with 150 successive trials consisting of pairs of manipulated prosodies (A) and asked to judge, within each pair, which sounded most interrogative (B). Patient responses in each trial were fitted with a 2-stage psychophysical model (C), consisting, first, of a prosodic template (or “internal representation”) to which sound stimuli are compared and, second, of a level of “internal noise” which controls how consistently this representation is applied to incoming stimuli. See main text for details about the model-fitting procedure. In this work, we estimate the two model parameters (representation and noise) for each patient individually and compare them with patient records to test their value as markers of receptive aprosodia.

**Figure 2.**
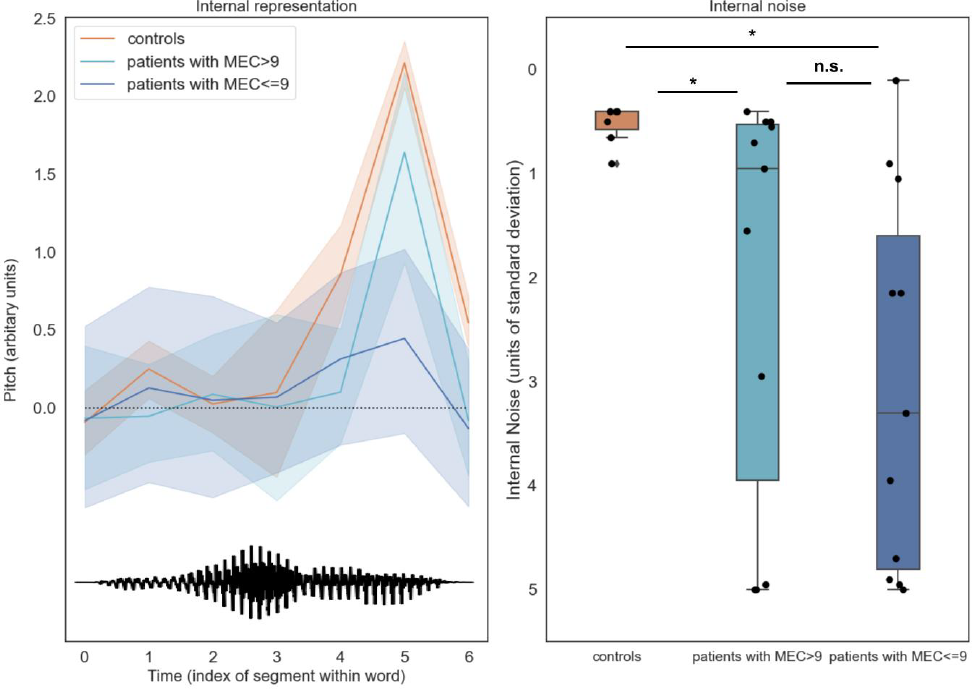
Patient parameters (internal representations and internal noise) estimated by reverse-correlation separate controls from patients above and below the pathological cut-off on the MEC prosody comprehension scale (9/12). Left: Internal representations of interrogative prosody computed from control group responses exhibited a typical final-rise contour, with a marked increase of pitch at the end of the second syllable. In contrast, patients’ internal representations had both lower amplitude and more variable shape across individuals. The bottom waveform illustrates the shape of the base sound used to generate stimuli (a male-recording of the word vraiment/really). **Right**: control participants were able to apply these representations remarkably consistently across trials, with internal noise values < 1 standard deviations of stimulus noise. In contrast, patients’ internal noise level were larger and more variable, and scaled with prosodic difficulties measured by MEC.

**Figure 3.**
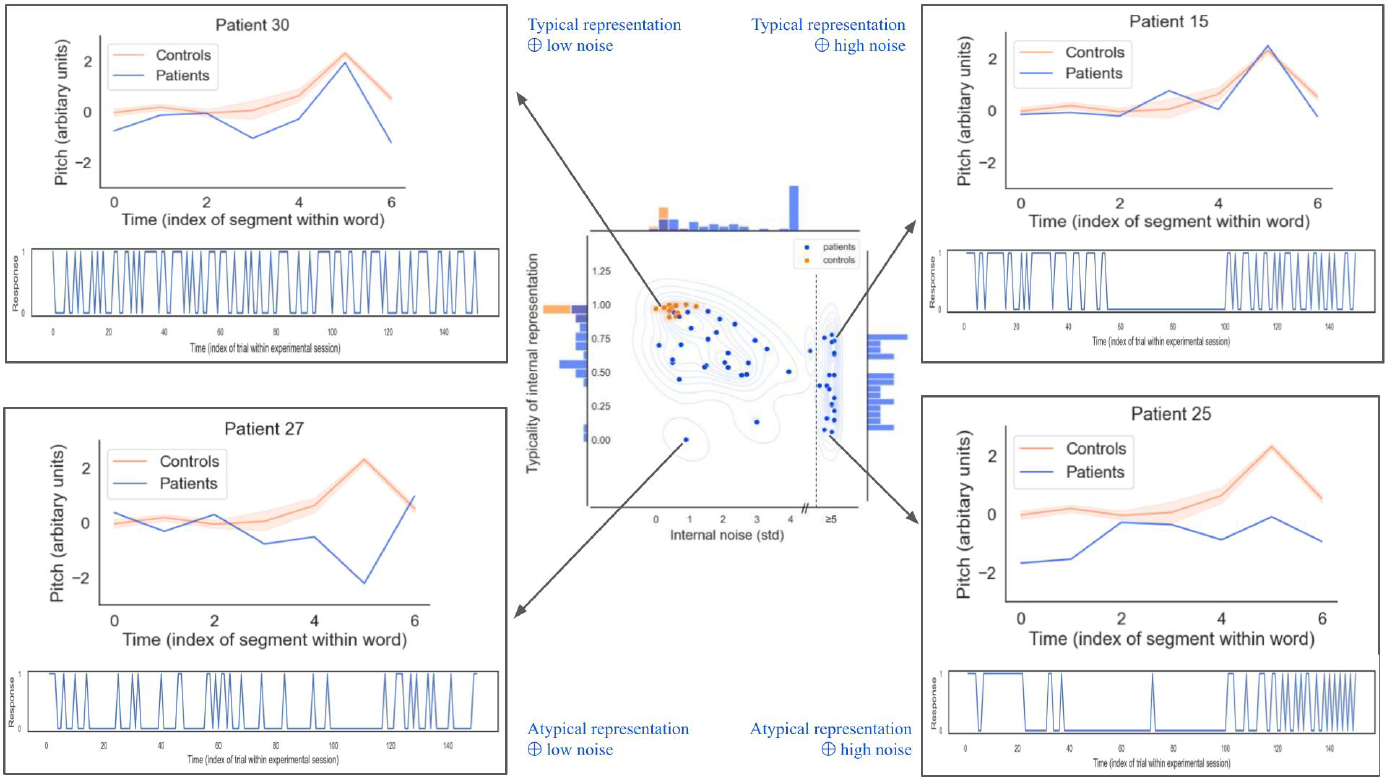
The representation+noise model captures a rich diversity of sensory/cognitive mechanisms underlying impairments of prosody processing after stroke. **Center**: Distribution of representation typicality and internal noise for controls and patients (considering all 4 sessions), overlaid with by kernel density estimate. Histograms on the marginal axes show univariate distributions for each variable in the patient group. **Corners**: Corner boxes show internal representations (top) and behavioral series of responses (bottom) for 4 illustrative patients. Patients in top corners have internal representations (blue) that are similar to controls (orange), but vary in amounts of internal noise (e.g. showing excessive response perseveration; top-right). Patients in bottom corners have atypical representations (blue), but some nevertheless retain healthy levels of internal noise (e.g., being normally consistent in wrongly expecting question phrases to decrease rather than increase in pitch; bottom-left). The estimation of internal noise was limited to the range [0; + 5std]; data points in the upper side of that range may either correspond to true internal noise values, or to larger values for which we could not provide an exact estimate, as illustrated here with a dotted line in the central panel (see Appendix for details).

### Reverse-correlation analysis

For each participant’s response data, we fitted a 2-stage psychophysical model consisting, first, of a prosodic template (or “internal representation”) to which sound stimuli are compared and, second, of a level of “internal noise” which controls how consistently this representation is applied to incoming stimuli (Figure 1).

Participants’ internal representations (a time x pitch representation of an ideally interrogative pitch contour) were computed using the classification image technique^7^: the mean pitch contour of the voices classified as non-interrogative was subtracted from the mean pitch contour of the voices classified as interrogative, and the resulting representation was normalized by dividing it by the sum of their absolute values. For each patient, we then quantified how similar their internal representation is to the average representation in the control group, by computing the mean squared error between the two representations, and used this “representation typicality” as a parameter to correlate with clinical measures. Representations for controls were computed using the same procedure, using only the first 150 trials of each session in order to match the number of trials seen by patients.

Participants’ internal noise (expressed in units of the standard deviation of stimulus noise) was inferred from response consistency and response bias across the repeated double-pass trials, using the simulation procedure of Neri (2010).^9^ In short, we computed an idealized participant model responding to repeated stimuli pairs of various sensory evidence, perturbed its response with additive gaussian noise (“internal noise”), and estimated the probability for that model to give the same response for identical trials (i.e. response consistency) and the probability of giving the first response option (i.e. response bias), for different standard deviations of that internal noise. For each participant, we then inverted that model and obtained the value of internal noise (by exhaustive search between 0 and +5 std) that minimized the error between the observed and predicted values for that participant’s consistency and bias. As in previous studies^9^, we estimated internal noise conservatively between [0; +5 std] in order to avoid unreliable estimates at large values, a known problem with double-pass procedures (see Appendix A). Internal noise values in the upper side of that range (e.g. illustrated in Figure 3 between 4.8 and 5) may either correspond to true internal noise values, or to larger values for which we could not provide an exact estimate.

Both of these analyses (internal representations and internal noise) were conducted using an open-source Python toolbox built for this purpose (https://github.com/neuro-team-femto/palin). See supplementary material (analysis code) for a complete description of the procedure.

### Statistical analysis

Group comparisons: because distributions of representation typicality and internal noise scores between patients and controls were non-normal, we compared population means using non-parametric (Mann-Whitney) independent sample t-tests.

Correlation with clinical measures: linear associations between representation typicality and internal noise, and clinical assessments (MEC, Prosody Comprehension, Prosody Repetition, Airtac2) met the homoskedasticity assumption and were therefore estimated using ordinary least-square regressions without robust (HC) norms, as these are considered to increase false positive rates when testing small samples^25^. In addition, because regression residuals were non-normal, we estimated statistical significance using bootstrapped confidence intervals. The analysis was implemented with the pymer.lm package.

### Ethics statement

The study was approved by *Comité de Protection des Personnes* CPP Ile-De-France V (ProsAVC, Decision of 22/07/2020). All participants provided informed consent.

## Results

Both measures extracted from the reverse-correlation procedure allowed separating patients from controls: internal representations of interrogative prosody computed from control group responses exhibited a typical final-rise contour^8^, with a marked increase of pitch at the end of the second syllable (Figure 2-left), and control participants were able to apply these representations remarkably consistently across trials, with internal noise values M=0.51 (SD=0.19) in the range of those typically observed for lower-level auditory and visual tasks^9^ (Figure 2-right). In contrast, patients’ internal representations had both lower amplitude (indicating less discriminative power) and more variable shape across individuals (Figure 2-left; see also Figure 3), and were applied with higher levels of internal noise (M=2.57, SD=1.93; Figure 2-right). The two groups differed statistically for both representation typicality: M=-0.28 [-0.39;-0.17], Mann-Whitley’s U(0.80)=25, p<0.000; and internal noise: M=2.06 [1.24; 2.86], U(-0.74)=134.00, p=0.004

Within the patient group, internal noise values (and, to a lower extent, representation typicality) were statistically associated with scores of the current gold standard for assessing deficits of prosody perception (MEC), demonstrating good concurrent validity. First, larger internal noise values were associated with lower (more severe) scores on the MEC prosody comprehension scale: noise: R^2^ = 0.19, β=-0.31 [-0.62; -0.08], t(20)=-2.22, p=.038. Representation typicality also improved with better scores, albeit non-statistically (R^2^ = 0.11, β=+0.03 [-0.003; +0.07], t(20)=1.57, p=.13). Second, both measures had also good symptom specificity, as strikingly neither correlated with the MEC score for prosody repetition (representation: R^2^=0.047, t(20)=-0.98, p=.34), while both MEC scores were themselves positively correlated (R^2^: 0.29, β=+0.32 [+0.08 – +0.56], t(20)=2.85, p=.01).

An oft-quoted limitation of the MEC instrument is its poor sensitivity, with patients above the pathological cut-off on the MEC prosody comprehension scale (9/12) still complaining of communication difficulties^5^. Interestingly, our measures allowed clear separation of this group of MEC-negative patients (N=12/22) and controls (N=12), both in terms of typicality of representation (M=-0.18 [-0.32 – -0.06], U(-0.68)=23.0, p=.005) and internal noise (M = 1.71 [0.71, 2.83], U(-0.7)=71,p=0.014).

Finally, to examine the convergent validity and specificity of internal representation and internal noise measures, we investigated whether they were statistically associated with other constructs linked to central deficits common in stroke rehabilitation. Expectedly, both measures were associated with difficulties discriminating tone intensity and tone duration, as measured by AIRTAC2 (representation: R^2^: 0.45, β=+0.04 [-0.001 – 0.062], t(11)= 2.97, p=.013; noise: R^2^: 0.35, β=-0.29 [-0.48; -0.06] t(11)=-2.43, p=.033). However, they were not associated with the patient’s capacity to detect rare auditory targets among distractors, as measured by LAMA (representation: R^2^: 0.015, t(10)=-0.38, p=.70; noise: R^2^: 0.002, t(10)=0.13, p=.89); or with the patient’s capacity to process musical melodies, as measured by MBEA scale and melody items (representation: R^2^: 0.06, t(11)=0.85, p=.41; noise: R^2^: 0.00, t(10)=-0.05, p=.96). Internal noise (but not representation typicality) was also found related to patients’ level of anxiety and depression, as measured by HADS (R^2^= 0.24; HADS: β=0.11 [0.015; 0.18], t(20)=2.50, p=.021).

## Discussion

In this report, we introduced a novel, simple psychophysical procedure which, by combining systematic digital manipulations of speech stimuli and reverse-correlation analysis, allows estimating the internal sensory representations that subtend how individual patients perceive speech prosody, as well as the level of internal noise that govern behavioral variability in how patients apply these representations in prosodic perceptual tasks.

Tested on a sample of N=22 right-hemisphere stroke survivors, our two proposed parameters of representation typicality and internal noise provide a promising alternative to the clinical gold standard for evaluating impairments of prosody processing (MEC). First, internal noise (and, to a lesser extent, internal representations) strongly associate with receptive, and not expressive, aprosodia measured by MEC within the patient group. Second, internal representations (and, to a lesser extent, internal noise) have better sensitivity than MEC for separating high-functioning patients from controls. Finally, both measures appear to have relatively good specificity with respect to non-prosody-related impairments of auditory attention and auditory processing, although internal noise was also found associated with mood disorders which, in our sample, were also predictors of MEC scores.

Taken together, the representation+noise model paints a simple yet potent portrait of the variety of sensory/cognitive mechanisms that can explain impairments of prosody processing after stroke: patients may differ from controls by having altered representations but a healthy level of internal noise (e.g., being *normally* consistent in *wrongly* expecting e.g. question phrases to decrease rather than increase in pitch - Figure 3-left); by having normal representations but abnormal levels of internal noise (e.g. showing excessive response perseveration and suboptimal executive control on top of otherwise *normal* sensory processing - Figure 3-right); or both.

By separating these different profiles of pathology, it is our hope that the representation+noise model will provide more effective and individualized therapeutic targets for rehabilitation of individuals with impaired speech prosody perception than existing measures. For example, patients with the highest levels of internal noise may benefit from therapies that focus on attentional and executive skills. In larger samples of patients, it will be also interesting to characterize the separate physiopathological determinants of abnormalities in either representations or noise, for instance in terms of different lesion locations or sizes. Finally, as the procedure is easy to dispense remotely and can be optimized to even shorter durations than the relatively light (~15min) procedure used here^27^, we are also interested in the potential prognosis value of measuring changing levels of representation typicality and noise longitudinally, along the course of rehabilitation.

## Supporting information

Supplementary -Text 1. Internal noise inference from double-pass experiments: Upper theoretical limit and limited sample size issues

## Data Availability

The data that support the findings of this study are available from the corresponding author, MV, upon reasonable request.

## Funding

This work was supported by grant from *Fondation pour l’Audition* (FPA RD 2021-12).

## Notes

### Competing Interest Statement

The authors have declared no competing interest.

### Clinical Protocols

https://clinicaltrials.gov/study/NCT05874011

### Funding Statement

This study was funded by Fondation pour l'Audition FPA-RD-2021-12

### Author Declarations

The study was approved by Comite de Protection des Personnes CPP Ile-De-France V (ProsAVC, Decision of 22/07/2020)

### Summary of Updates

There was a mistake in the title, it's " Separating representational and noise components of speech prosody perception after brain stroke"

