## Supplementary -Text 1. Internal noise inference from double-pass experiments: Upper theoretical limit and limited sample size issues for "Separating representational and noise components of speech prosody perception after stroke"

In this work, we model the perceptual decisions made by brain-stroke survivors regarding classifying words as interrogative/not, based on their pitch contour. To do this, we use the experimental paradigm of reverse-correlation to extract parameters of a model composed of (1) an internal "representation", or prosodic template (also called psychophysical kernel, or classification image in the psychophysical literature; Murray, 2011) and (2) a measure of "internal noise" which is inferred from response consistency and response bias across the repeated double-pass trials, using the simulation procedure of Neri (2010). Because of technicalities in this procedure, and the limited number of double-pass trials that can be presented to patients, our methodology lacks robustness to estimate larger values of internal noise (measured in units of stimulus noise standard deviation). In the present study, we conservatively limited the search for internal noise values between [0; +5 std], and noted that values in the upper side of that range may correspond to either larger (and probably pathological) values of internal noise, or to participant behavior which did not conform well to the representation + noise model, both of which would be clinically relevant. The goal of this appendix is to give further details about the limits of the double-pass procedure for estimating large values of internal noise, using computational simulations.

The internal noise inference procedure aims to convert data obtained in double-pass experiments under the form ($P_a$, $P_{int1}$), where $P_a$ stands for the percentage of agreement, i.e. giving the same response, over repeated trials, and $P_{int1}$ stands for the probability of selecting the first response category (also called first "interval" in psychophysics)over the set of repeated trials, into levels of internal noise (hereafter IN, expressed in units of external noise standard deviation). This conversion from ($P_a$, $P_{int1}$) to IN is done by simulating an ideal participant responding to repeated stimuli pairs with various levels of IN, estimating the resulting ($P_a$, $P_{int1}$) of the model, and then invert the model by searching for the value of IN that minimizes the error between the observed and the predicted values for ($P_a$, $P_{int1}$). This procedure has a lot of theoretical advantages, namely that it  provides results that do not depend on the experimental design and levels of performance, and that can be directly compared across modalities. However, the ($P_a$, $P_{int1}$) to IN  conversion is only possible and valid in a restricted region of the full ($P_a$, $P_{int1}$) space. In practice, most studies that use this inference approach only consider inferred IN values within a limited range of [0.2 – 5 units of ext. noise SD] (Neri, 2010).

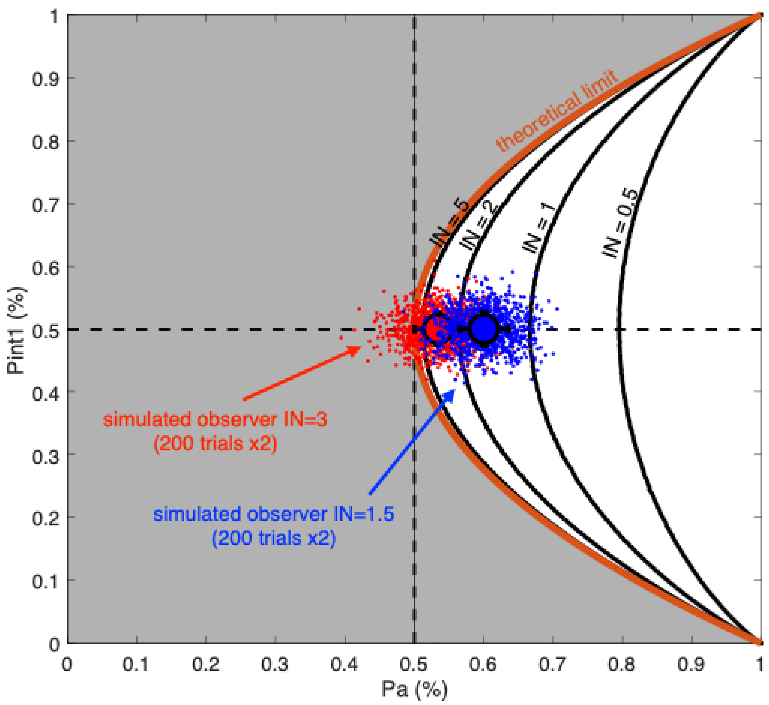


***Supplementary Figure 1*** *– Limits of the internal noise inference approach illustrated with data from double-pass, two-interval experiments, presented in the* ($P_a$, $P_{int1}$) *space (*$P_a$*: probability of agreement over repeated trials;* $P_{int1}$*: probability of selecting the first response category across repeated trials). The orange line shows the upper theoretical limit for an infinite level of internal noise (i.e. responding randomly); the black lines show several iso-internal noise curves (levels of 0.5, 1, 2 and 5 units of external noise SD) generated from computational simulations. In order to illustrate the limits and the inherent sources of variability in the inference procedure when using empirical data obtained with a limited number of trials, we superimposed hypothetical data obtained by simulating two model observers (with no response bias, each point in the space corresponds to one simulated double-pass experiment of 200 trials x2; the model was run to simulate 1000 experiments) with plausible levels of internal noise (1.5 or 3 units of external noise SD) performing double-pass experiments. ( For each run, the internal noise inference algorithm will try to infer (maximum likelihood procedure) the closest internal noise level from*

*the position of the ($P_a$, $P_{int1}$) that best fits the observed data. When empirical data ($P_a$, $P_{int1}$) fall above (or too close to) the theoretical limit of infinite internal noise, the internal noise inference approach cannot be used reliably. This can occur because of a limited number of trials or more fundamentally because the sensory or cognitive process in the experiment does not align with the underlying SDT-based hypothesis of the model (see the text for details).*

The introduction of an upper limit for inferring internal noise comes from the fact that empirical values of ($P_a$, $P_{int1}$) can fall close to, or even above, the theoretical limit between the two quantities described by the curve $P_a = P_{int1}$^2 + (1- $P_{int1}$)^2, which correspond to responding at random on each trial: agreement on a given trial is either because the model responds 1 with probability $P_{int1}$ on both trials, an event with probability $P_{int1}$ x $P_{int1}$, or because the model responds 2, wth probability 1-$P_{int1}$ on both trials, an event with probability (1-$P_{int1}$)*(1-$P_{int1}$). The region of the ($P_a$, $P_{int1}$) space in which internal noise values cannot be inferred is colored in grey in **Supplementary Figure 1**. It is important to distinguish between two cases in which this situation can happen:

(1) This can happen because, in practice, double-pass experiments are run with a limited number of repeated trials. For instance, an unbiased observer can lead to a percentage of agreement falling below 50% – which would not be possible with an infinite number of trials. As illustrated in **Supplementary Figure 1**, this case becomes more frequent for observers with higher levels of internal noise, because the theoretical (Pa, Pint1) point that would be obtained given an infinite number of repeated trials comes closer to the theoretical limit of infinite internal noise. Our simulations show that in a double-pass experiment with 200 trials repeated twice, estimations above the theoretical limit occur in less than 1% of simulations for a theoretical observer with IN =1.5 but in ~15% of simulations for a theoretical observer with IN =3. Put differently, the number of repeated trials that would be necessary to estimate large values of IN reliably (i.e. with the same confidence interval reached here for values of INs limited to the [1-5] std range) would be prohibitively large, leading to experiments of impractical duration for fatigable patients in our clinical context.

(2) Estimated values of (Pa, Pint1) that fall above the theoretical limit may also happen not because of an estimation problem linked to limited sample size, but because the true value of (Pa, Pint1) is indeed located in that region. This can be the case if the patient's cognitive process generating double-pass data is not fully consistent with the ideal observer model used to estimate the IN value that best simulates their behavior. The observer model used to estimate internal noise (also called signal detection theory, or SDT, model) assumes constant behavior throughout the experiment, and does not account for many of non-regularities that may be observed in healthy participants (e.g. learning, criterion shirt between passes) or patients (e.g. fluctuations of attention, perseverations). This aspect was notably discussed by Spilioti et al. (2016) when trying to assess internal noise in fishes using a double-pass procedure. They found several cases of collected double-pass data falling outside of the theoretical region, bringing the question of whether the fish responses were related to the characteristics of the stimuli, a prerequisite before deploying the approach. We believe that this limitation is especially relevant when working with atypical populations of patients who can produce extreme behaviors not generally observed in lab-based experiments with healthy individuals. For instance, perseverations could in an extreme cases produce a large number of responses on interval #1 on the first pass and interval #2 on the second pass, producing an average Pint1 of ~50% but a Pa close to 0%.

It is impossible to distinguish between a limited sample size issue and the incompatibility of the collected data with the SDT-model. It is nevertheless interesting to note that > 10% even for the double-pass data collected in low-level psychophysical experiments reviewed in Neri (2010) fall above the limit and from which an internal noise level could therefore not be inferred. In practice, the upper limit has often been empirically defined at *IN*=5. As illustrated with our simulations, the number of trials required for a given *IN* precision increases exponentially with *IN*, and it can be noticed that above 5, the iso-internal noise curves become descriptively so close to each other that the required number of trials to robustly distinguish between different internal noise levels makes the inference approach experimentally impractical.

In the present work, we estimate patient IN by restricting search between [0-5] std. It is likely that data points falling in the upper side of this range correspond to patients with either theoretically larger values of internal noise, but which our limited number of repeated trials would not allow to robustly estimate, or whose behavior departs more severely from the constant-behavior SDT model. While in our data this relatively simple description of IN already correlates with aprosodia symptoms and allow to separate patients with e.g. correct but inconsistent representations from more severe patients with both incorrect and inconsistent representations (main text Figure 3), our study calls for further methodological development on the assessment of internal noise in atypical populations, and/or of other more sophisticated observer models that incorporate other pathological variables of interest, such as the probability to enter episodes of perseveration.
